# Modeling Covid-19 dynamics for real-time estimates and projections: an application to Albanian data

**DOI:** 10.1101/2020.03.20.20038141

**Authors:** Erida Gjini

## Abstract

The SARS-CoV-2 epidemic is one of the biggest challenges healthcare systems worldwide have ever had to face. To curb transmission many countries have adopted social distancing measures and travel restrictions. Estimating the effect of these measures in each context is challenging and requires mathematical models of the transmission dynamics. Projections for the future course of the epidemic strongly rely on model predictions and accurate representation of real-time data as they accumulate. Here I develop an SEIR modeling framework for Covid-19, to evaluate reported cases and fatalities, and to enable forecasting using evidence-based Bayesian parameter estimation. This Bayesian framework offers a tool to parametrize real-time dynamics of Covid-19 cases, and explore the effect of control as it unfolds in any setting. I apply the model to Covid-19 data from Albania, where drastic control measures were put in place already on the day of the first confirmed case. Evaluating the dynamics of reported cases 9-31 March 2020, I estimate parameters and make preliminary projections. Three weeks into the measures, Albanian data already indicate a strong signature of more than 40% transmission reduction, and lend support to a progressively increasing effect of control measures rather than a static one. In the Albanian setting, the model and data match well, projecting the peak of the outbreak may be around 5-15 April, and be contained within 300 active confirmed cases if control continues with the same trend. This framework can be used to understand the quantitative effects of different control measures in real-time, and inform adaptive intervention for success in other settings.

## Current data and situation

Albania reported the first confirmed case of coronavirus disease (COVID-19) on the 9th of March 2020. Given very close travel connections with Italy, where the epidemic grew very fast over the month of February, the concern over a major similar outbreak in Albania was significant. Immediately following the first case confirmation, country-wide measures were taken to drastically reduce transmission and contain the spread of infection. These included university and school closures, suspension of public gatherings, closure of bars and cafes, travel restrictions, and gradually a lockdown of the population. The parameters governing the epidemic course of COVID-19 in Albania remain unknown. The first week after the first detected case, official numbers indicated case counts amounting to a total 59 confirmed cases on March 18, with two fatalities (Albanian Institute of Public Health). The initial exponential growth rate of the number of cases in Albania was *r* = 0.17 during the first week, coming from a doubling time of 4 days (1), but this has slowed down from March 14 onward. It is not known what will be the trajectory of the epidemic in this country. Following the diagnosis of the first cases, the borders with Italy, Greece, Kosovo were rapidly closed and many flights to other European destinations were limited or cancelled.

The aim of this study is to provide a framework for interpreting and forecasting the real-time course of the epidemic as it unfolds in a given setting. I use the Albanian data as an illustration for the application of the model. With the model and Bayesian estimation, continuously-updating values for the parameters, underlying the epidemic course of COVID-19, can be obtained, and continuously-updating projections. Based on the model application to Albania, the parameter estimates and their associated uncertainty, suggest clear positive effects of control measures and make precise quantitative projections about the magnitude and timing of the peak of this outbreak in Albania.

## Mathematical model

There are currently many efforts to understand Covid-19 dynamics around the world using mathematical models (2; 3; 4; 5). The mathematical model I use here is based on SEIR dynamics (Figure 1, simplifying the model by (4) to a version without age structure. We model the number of **susceptible** individuals *S*, **exposed** individuals *E* whose transmission may be considered negligible, **infected symptomatic** *I*_*S*_ and **infected asymptomatic** *I*_*A*_ individuals, as well as **fatalities** *F* and **recovered** individuals *R*. The total population size is given by *N* = *S* +*E*+*I*_*S*_ +*I*_*A*_+*R*+*F*. At the beginning of the epidemic we have *S*(0) *≈ N*.

**Figure 1:**
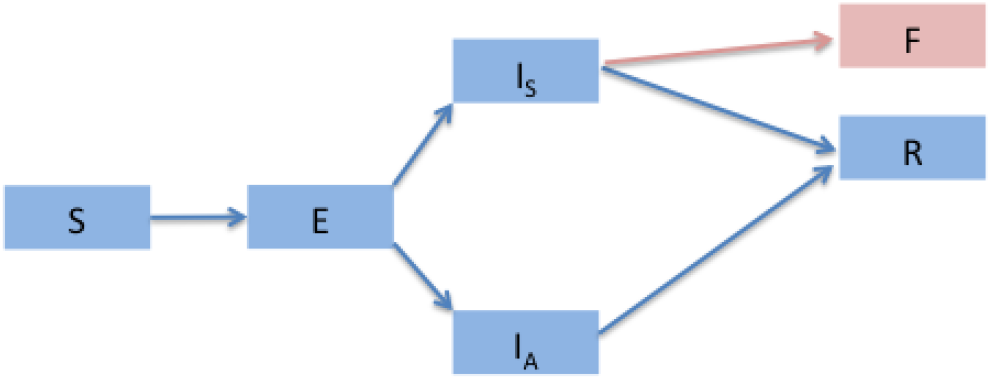
Epidemiological model schema. Transmission dynamics from Susceptible (*S*) to Exposed (*E*) to Infected symptomatic (*I*_*S*_) or Infected Asymptomatic (*I*_*A*_) and then finally to Recovered (*R*) or Fatality (*F*) cases. The transitions between compartments are modeled with ordinary differential equations. I follow the structure in (4) but without age structure. Confirmed cases in the data are assumed to reflect the magnitude of the symptomatic compartment in the model.

The ordinary differential equations describing the transmission dynamics are as follows:

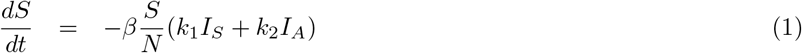

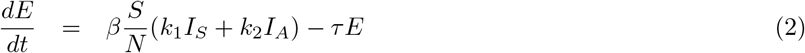

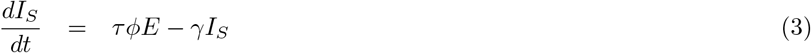

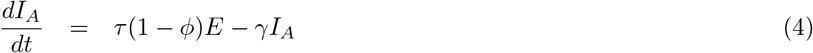

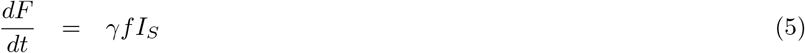

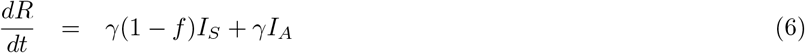

For the parameters, average values were assumed based on the existing literature on COVID-19 (6; 7; 4). The basic reproduction number *R*_0_ for COVID-19 transmission, denoting the number of new infections generated from a typical infected individual during the period of infectiousness, is expected to be between 1.7 and 2.6, (2), and is the key target of control measures. The incubation time from exposure to infection, informing the progression rate *τ*, has been shown to be around 6.5 days (median), and up to 14 days (8). Exposed individuals are assumed to not be infectious. After progressing to infection, a fraction of individuals are symptomatic given by *Φ*, and 1 *− Φ* are asymptomatic. The duration of infection from infected people in Wuhan has been observed to be around 11 days (7), motivating my assumption of *γ* = 1/11 in the model, but larger and lower values have also been reported such as 20-30 days infectious period (9; 6) and 3.48 days (3). In Albania, until March 28, more than 30 infected individuals have recovered, with typical viral shedding from onset of symptoms for about 6 days.

The model assumes symptomatic individuals can progress to recovery or death with relative probability 1 *− f* and *f*. It is assumed that asymptomatic individuals only experience mild infection and progress to recovery. Asymptomatic individuals transmit less infection, scaling their contribution by a factor *k*_2_ (*k*_2_ *<* 1).

### Effect of control

The control measures in Albania started to be implemented very quickly, remarkably starting at the first day of case detection, and scaling up to country-wide quarantine by March 14, 5 days afterwards. To implement the effect of control measures in the model, I included a time-varying (decreasing) *β*, assuming this started to be effective at day *T*_*in*_ after the first detected case:

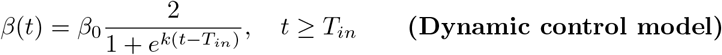

where the parameter *k* reflects the speed of reduction in transmission. Another way to model reduction in transmission is to assume an instantaneous drop in global transmission rate, by a factor *c* (constant) at time *T*_*in*_:

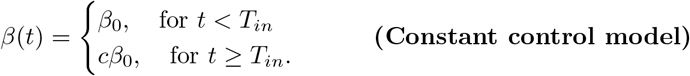

Both of these modes of control are fitted to the data, and their quality of fit is compared to verify which mode of control (dynamic or static) is more consistent with the dynamics observed.

### Bayesian inference

I used a Bayesian framework to estimate parameters fitting the model to data. We obtain the posterior distributions for *θ* as:

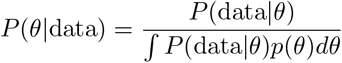

I used flat priors *p*(*θ*) for all parameters. The likelihood *P* (data *θ*) is given by a normal distribution for the mean-squared error between the data and the model. We take into account the data *d*_*i*_, *f*_*i*_ denoting number of active confirmed cases and cumulative fatalities for each time *t*_*i*_, and compare them with model predictions for *I*_*s*_(*t*_*i*_) and *F* (*t*_*i*_). For simplicity, I do not model the sampling process (e.g. Poisson) in this basic version.

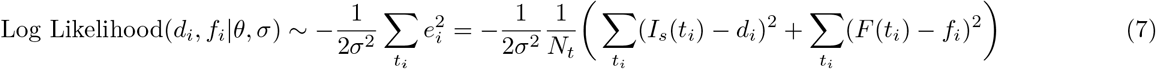

## Fitting initial dynamics of confirmed cases

I assumed that the number of confirmed (documented) cases reported reflects the number of symptomatic cases in the model, and that fatalities due to the disease are all reflected in the deaths observed among confirmed cases. Thus I fitted theoretical model trajectories (*I*_*s*_(*t*), *F* (*t*)) to the observed data, using the mcmcstat package (10), under a Bayesian framework. The parameters estimated were *E*_0_, *β*_0_, *f* and *k* or *c* depending on the assumed mode of control (dynamic/constant). For the other parameters, I assumed fixed values as in Table 1. In this model, the basic reproduction number is given by:

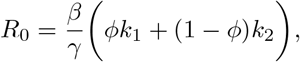

while the effective reproduction number *R*(*t*) accounts for dynamic changes in *β*(*t*) due to intervention measures.

**Table 1:** Parameter values used for model calibration to the Albanian setting.

| Parameter | Meaning | Value | Reference |
| --- | --- | --- | --- |
| $\beta_0$ | Baseline transmission rate | *Estimated* (per day) | $\beta_0 \in [0.2, 0.8]$ |
| $k_1$ | Scaling factor for infectiousness of symptomatics | 1 | (3) |
| $k_2$ | Scaling factor for infectiousness of asymptomatics | 0.55 | (3) |
| $\gamma$ | Clearance rate | 1/11 (per day) | (6) (7) |
| $\tau$ | Progression rate to infection | 1/6.5 (per day) | (6) (8) |
| $\phi$ | Probability of symptomatic cases | 50 – 60% | (4) (7) |
| $k$ | Rate of reduction in transmission via dynamic measures | *Estimated* | $k \in [0, 0.2]$ |
| $c$ | Relative reduction in transmission (instantaneous control) | *Estimated* | $c \in [0, 1]$ |
| $f$ | Case-fatality ratio | *Estimated* | $f \in [0, 0.5]$ |
| $S(0)$ | Number of susceptible population at day 0 | $N = 2.87 \times 10^6$ | Albania |
| $I_s(0)$ | Number of symptomatic cases confirmed at day 0 | 2 | Albania ISHP |
| $E(0)$ | Number of exposed individuals at day 0 | *Estimated* | $E_0 \in [1, 5000]$ |
| $I_A(0)$ | Number of asymptomatic individuals at day 0 | Assumed | $I_A(0) = 0$ |
| $F(0)$ | Number of fatalities at day 0 | Assumed | $F(0) = 0$ |
| $R(0)$ | Number of recovered individuals at day 0 | Assumed | $R(0) = 0$ |

Figure 2, A to C, shows model predictions of reported cases generated using the best-fitting model parameter estimates. The model fit captures the range of observed cases well, from March 2, 2020 up to March 28 2020. In addition, the best-fitting model yields posterior credible intervals for the future course of the novel coronavirus (COVID-19) outbreak in Albania. The Bayesian framework yields estimates for the best-fitting parameters as well as their posterior distributions (Figure 3). The estimate for the median crude-case fatality ratio is 7%, 95%CI(0.02,0.13), similar to such crude estimates obtained in early dynamics in Wuhan (3), but higher than the 1-2% true case-fatality ratio expected if all (symptomatic and asymptomatic) cases are confirmed (7).

**Figure 2:**
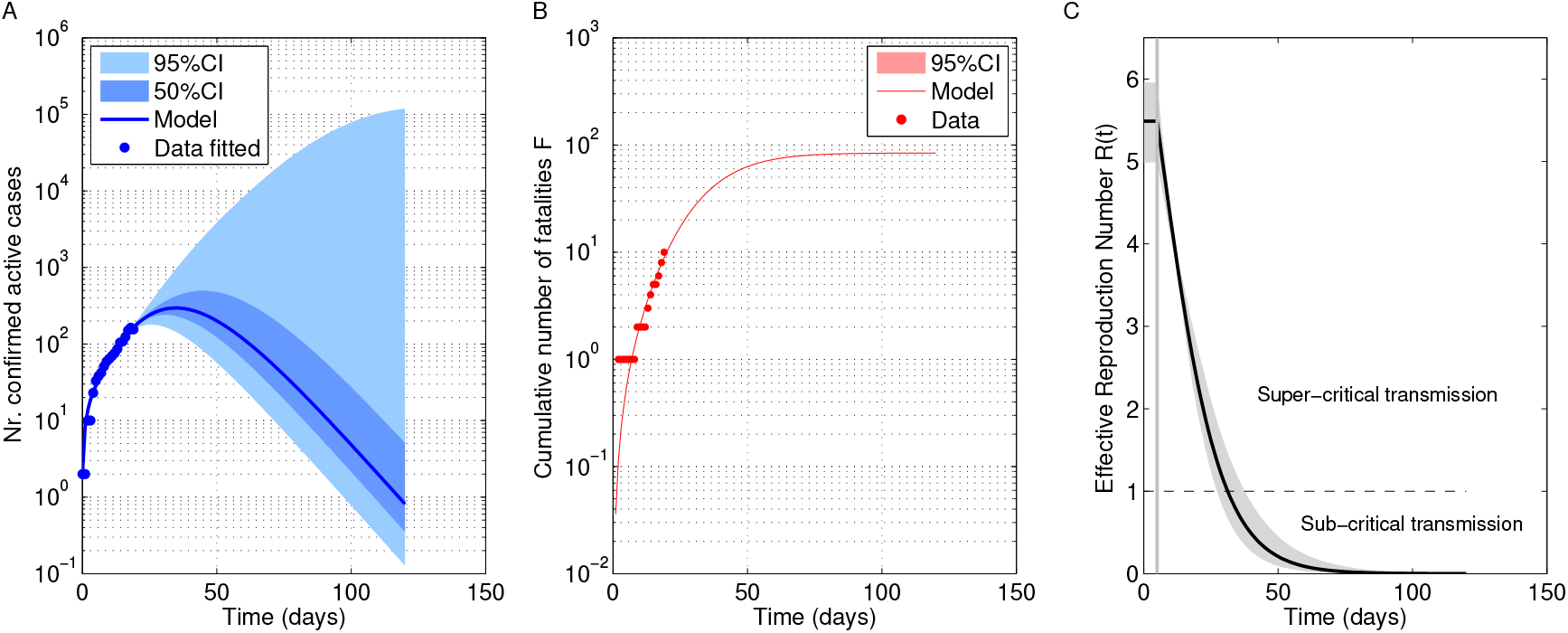
Model fit to Albanian data from March 9 to 28. A. Model prediction with best-fitting parameters for the number of confirmed cases (*I*_*S*_(*t*) in the model). The lighter shaded region gives the 95%credible envelopes with 100 simulations parametrized from random samples of the posterior distribution. The darker-shaded region gives the 50% credible envelope. B. Model prediction for the cumulative number of fatalities. C. Model prediction for the rate of reduction in transmission consistent with data up to March 28, 2020.

**Figure 3:**
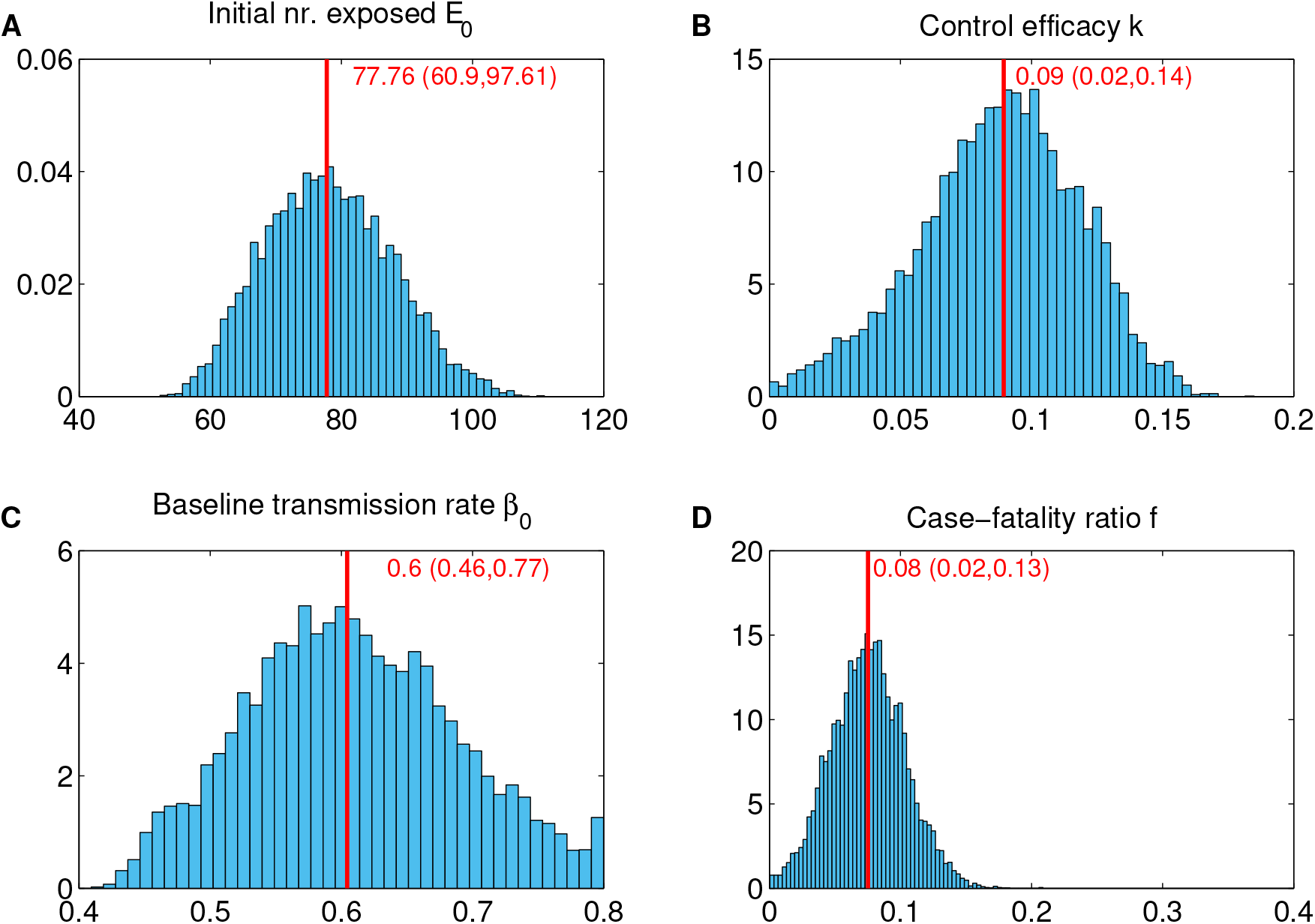
Posterior distributions of parameters using data from March 9 to 28. The mean-squared error between model and data is assumed normally-distributed with mean 0 and standard deviation *σ*. I used flat priors for all parameters. Two Monte Carlo Markov Chains were run for 5000 iterations until burn-in. Following convergence, another 10000 iterations were run to obtain the posterior distributions for all parameters (shown here) and to compute credible envelopes.

## Estimated *R*_0_ and control efficacy

The median estimate of the basic reproductive number (*R*_0_) at the beginning of the outbreak is 5.45, 95%CI (4.15, 6.95), about double the estimates from early dynamics in the literature (2; 3). The median estimate of the efficacy of dynamic control measures, assuming they were effective 5-days after the first confirmed case (*T*_*in*_ = 5) is 0.09, 95%CI (0.02,0.14). This implies that by the first week of control, transmission was reduced by about 30%.

## Estimated initial number of exposed cases

The initial seeding of the epidemic in Albania is unknown, but for the initial number of exposed cases *E*_0_, the model estimates a value of 78, 95%CI (61,98). This seems relatively small given the strength of travel connections with Italy. Yet, when we take into account the actual numbers, we obtain an independent estimate that is very close to this model-based estimate.

### An independent estimate of imported cases from Italy

A big unknown is the initial number of imported cases from Italy to Albania during the month of the epidemic outbreak in Italy (8th of February until 8th of March), just prior to the first case detected in Albania. Using the data available from INSTAT (11), about the yearly travel fluxes between Albania and Italy via air (1.7 million passengers) and by sea (1.6 million passengers), we obtain a total of 1.65 million arriving passengers on average from Italy to Albania in one year. This translates to 0.13 million passengers in one month. Thus, during the month prior to the first case in Albania, about 130.000 thousand passengers are expected to have arrived to Albania from Italy via air and sea. To compute the prevalence of infection expected in such passengers, we must account for the exponential disease dynamics in Italy during the same period. The prevalence of infection in travelers arriving to Albania from Italy can be assumed to be roughly the same as the prevalence of infection in the Italian population itself during this period (12). I used the publicly available data for the epidemics in Italy (1), starting with 3 cases in February 8 and counting 5883 cases on the 8th of March, following an exponential growth at rate *r* = 0.25 (13).

Using the population of Italy, 60.48 million, as denominator, this leads to:

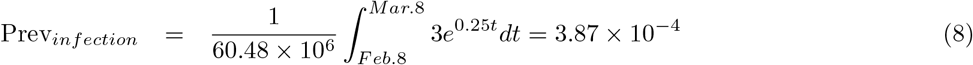

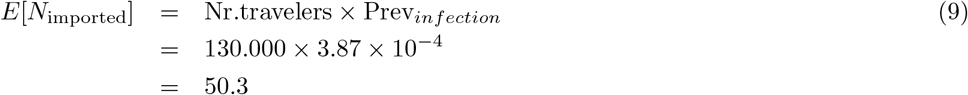

is the number of expected imported Covid-19 cases during one month in Albania from Italy, computed from travel statistics and epidemics in Italy. This number (*≈* 50) is close to the number exposed estimated with the model (*E*_0_ = 78) under assumed symptomatic percentage of 60%, incubation period of 6.5 days, and infectious period of 11 days. This match adds confidence to support the model results.

### Epidemic projections based on dynamics 9-28 March

Quantitatively, if the control measures continue with the same rhythm, this model with its best-fitting parameters, based on the data up to March 28, 2020, predicts that the peak of the epidemic in Albania will be reached on day 36 after the first detected case, with 50%CI (32,45) and 95% CI(26, 121). The magnitude of the number of active confirmed cases at the peak will be around 302 with 50% CI(242,498) and 95%CI(180,1.1 *×* 10^5^), thus still with substantial uncertainty. The total number of Covid-19 cases, including those documented/confirmed and those not-confirmed (asymptomatic) will be 1903 with 50%CI (1386,3674) and 95% CI (930, 5.4 *×* 10^5^). While all these predictions are to be taken with caution, since we don’t have yet a clear final signature of drop in cases, they indicate some reason for optimism in the possibilities of limited epidemic size of SARS-Cov2 in Albania. If the effect of control measures continues with this trend, the epidemic projections for Albania suggested by the model appear manageable. Under constant control instead, the data suggest that current efforts have reduced transmission by 40% (see Table 2), but if this remains at this level, it is not sufficient to limit the number of cases in the short term (see Supplementary Figures S1-S3).

**Table 2:** Model-based estimates for parameter values in the Albanian setting. These estimates were obtained using the confirmed case data 9-28 March 2020, and assuming the control was effective 5 days after the first detected case.

| Parameter | Meaning | Estimate: Median (95%CI) |
| --- | --- | --- |
| $\beta_0$ | Baseline transmission rate | 0.60(0.46, 0.77) |
| $k$ | Rate of reduction in transmission (dynamic control) | 0.09(0.02, 0.14) |
| $c$ | Relative reduction in transmission (constant control) | 0.60(0.50, 0.93) |
| $f$ | Crude case-fatality ratio | 0.07(0.02, 0.13) |
| $E_0$ | Number of exposed individuals at day 0 | 77.76(60.9097.6) |
| $R_0$ | Basic reproduction number | 5.45(4.15, 6.95) |

## Updating projections with data in real-time and model comparison

The advantage of this framework is that new data arriving in real-time can update model estimates and subsequent projections. In the case of Albanian data, adding the case observations for the next 3 days (March 29-31), narrows dramatically the credible intervals of the future course of the epidemic (see Figure 5) with little change in the posterior parameter estimates (Figure S4). The model-predictions for the quantitative aspects of the epidemic are also narrowed substantially (Figure S5). In particular, the peak of the outbreak is expected to occur between 5-15 April (Fig.S5C), and not surpass 300 active confirmed cases (Fig.S5B), if control continues with this trend. Comparing the model fits to data March 9-31 for the dynamic control and constant control model, we observe a smaller error for the dynamic control than the constant control model (Figure 6). This is further confirmed from analysis of the deviance information criterion DIC (14), where we find that *DIC*_*dynamic*_ = 4447 for the dynamic model, and *DIC*_*constant*_ = 27283 for the constant model. Typically models with smaller DIC are favoured. Thus, the data strongly suggest that the effect of control measures on transmission in Albania has been dynamic, increasing over time.

**Figure 4:**
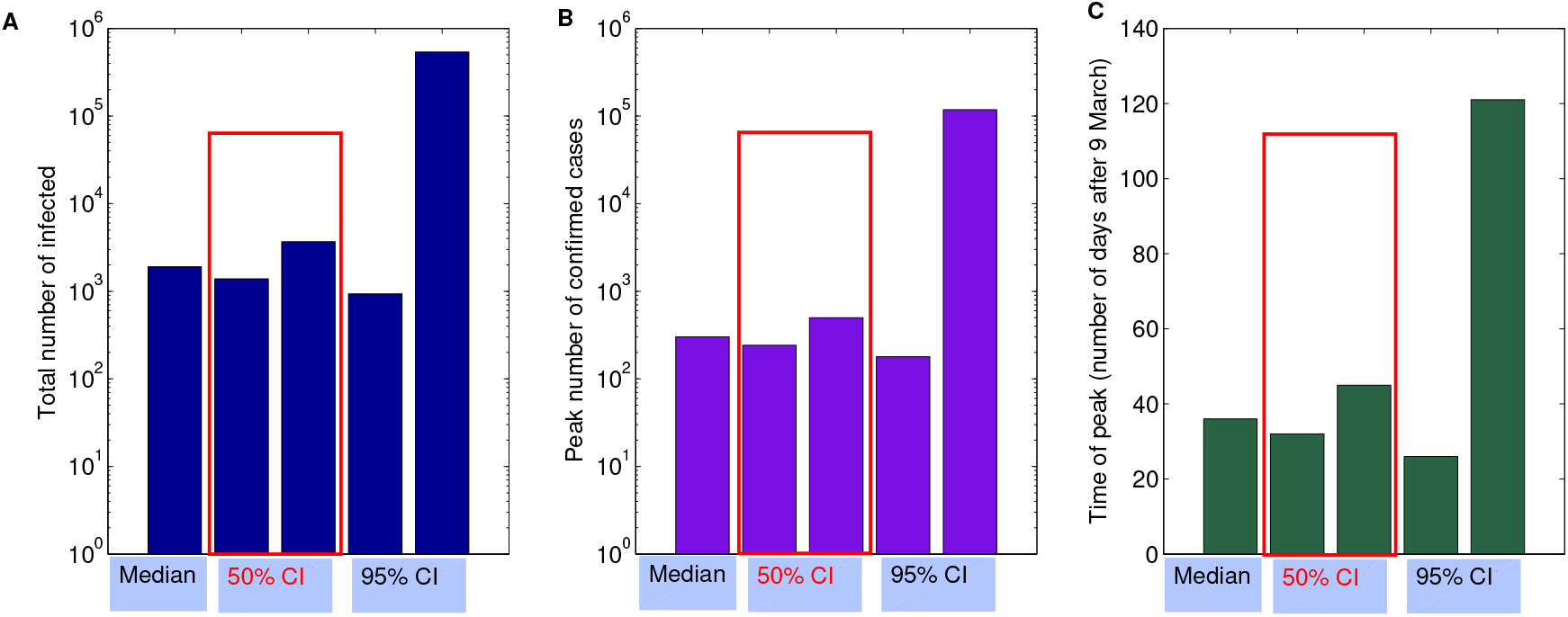
Epidemic projections based on dynamics 9-28 March. Using the model with best-fitting parameters and uncertainty (Figure 2), the dynamics are simulated forward in time to compute statistics about the epidemic course of Covid-19 in Albania. A. Total number of infected. The projection suggests a median number of total number of infected individuals about 1500. B. Peak of confirmed cases. The model suggests a median of about 300 confirmed active cases. C. Time of peak of the outbreak. The model predicts the peak will be reached around mid-April 2020.

**Figure 5:**
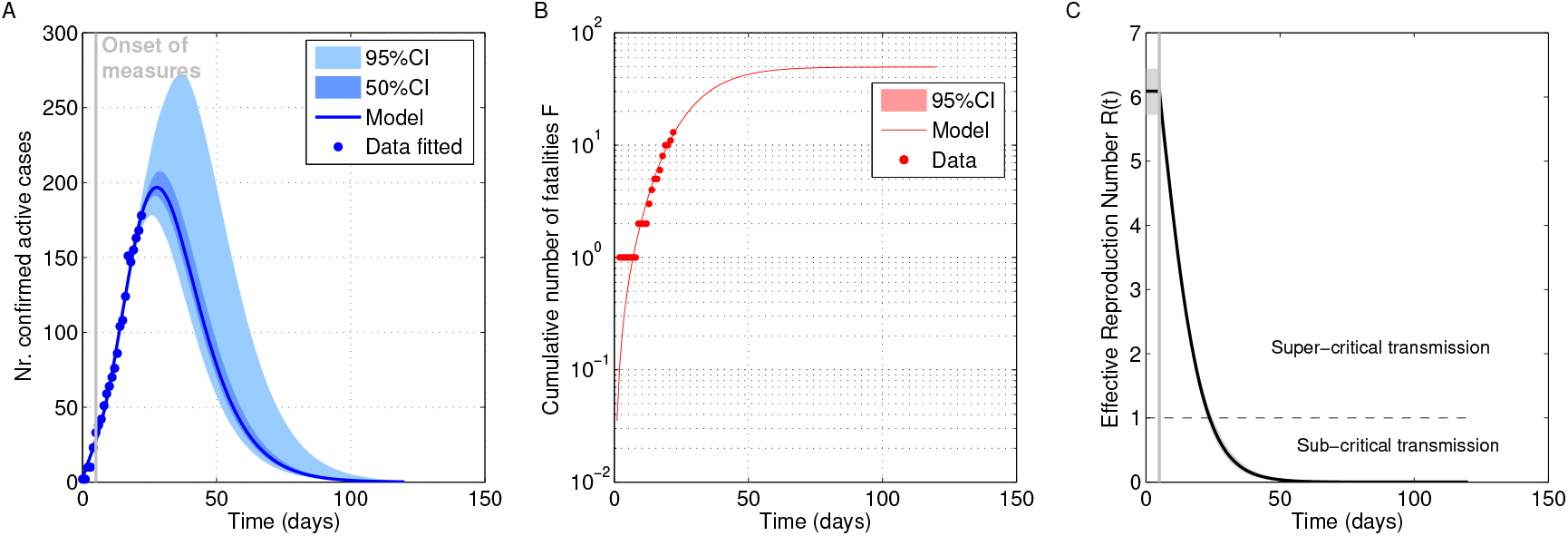
Model fit and projections based on dynamics 9-31 March. Feeding the model with more data substantially enhances model predictions, reducing the uncertainty, and suggesting a small size epidemic in Albania.

**Figure 6:**
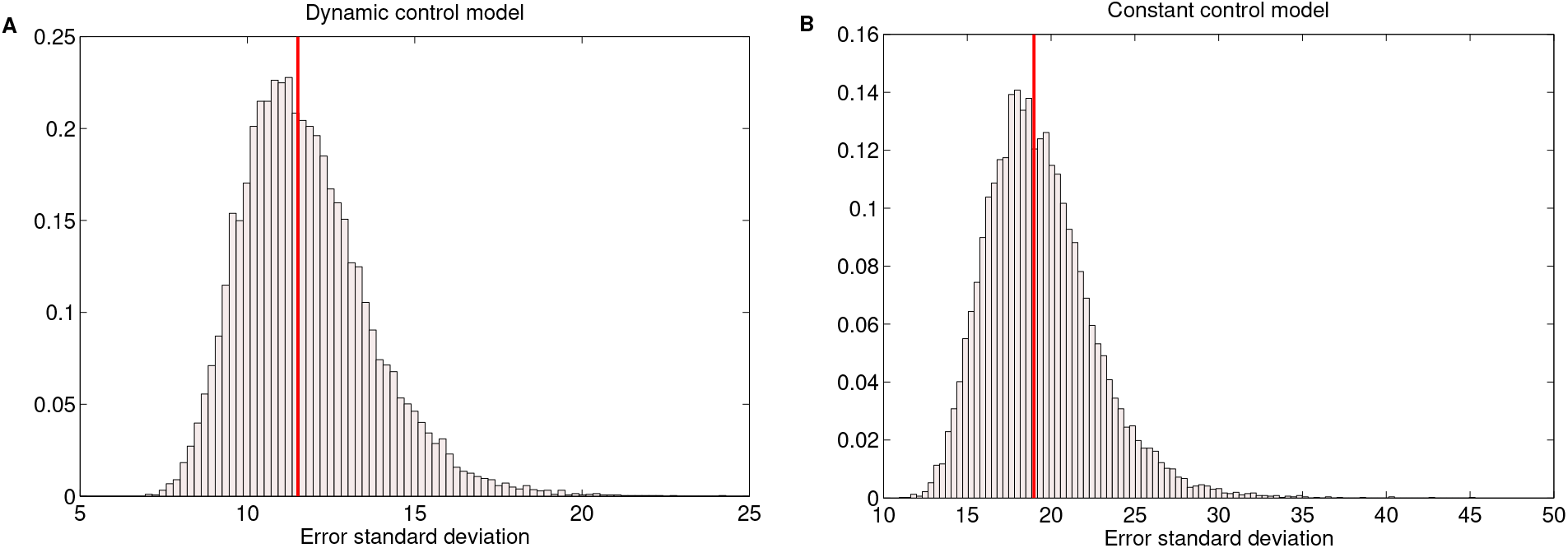
Error comparison for the dynamic control and constant control model, based on larger dataset 9-31 March. The data are strongly suggestive of the dynamic effect of control measures in Albania, supporting a progressively decreasing *β*(*t*) since March 14 onwards.

## Discussion

This is a simple model embedded in a Bayesian framework, to fit real-time data of early COVID-19 dynamics, estimate parameters and generate predictions, accounting for intrinsic uncertainty. The model is generic and can be applied to any setting. I show that it captures well the data from March 9 to March 28 in Albania, and suggests that the effects of control are already tangible in the dynamics. A dynamic control put in place almost immediately after the detection of the first case, has had an effect of reducing the original transmission, already by 30% in the first week of implementation. The projections are based on maintaining control from the onset of intervention, *T*_*in*_, throughout the period simulated.

The model assumes homogeneous mixing of all individuals, which is of course an approximation. There are clear social networks and heterogeneities between regions, age classes, socio-demographic groups, especially households, that the model is not considering. Stochasticity, for example in person-person variation for number of new infections generated, considered by other studies (2) could be important also in this context, where the numbers of infected reported are relatively low.

The model assumes all symptomatic infections are detected. If confirmed cases only reflect a proportion of the true symptomatic cases, then the effective symptomatic proportion the model would ‘see’ would be lower, and slightly different model parameters would result.

An *R*_0_ so high, of the order of 5, estimated by the early dynamics of confirmed cases in Albania, may be related to superspreaders having dominated the early transmission events in the epidemic in this country, or related to big public gatherings and festivities at national level (political meetings, International Women’s day, etc.) just preceding the first confirmed case. Other studies have also shown that mean *R*_0_ ranges from 2.24 (95%CI: 1.96-2.55) to 3.58 (95%CI: 2.89-4.39) (15), and is very sensitive to the reporting rate. If the reporting rate is increased by 8-fold, the estimate is expected to approach the more widely agreed values in the literature (2; 3). So far the model does not take into account reporting rate, but it is likely that Covid-19 cases in Albania may be under-reported, due to the low number of overall tests performed, compared to the total population size.

Although at first sight it may seem that the model does not capture the specific isolation of contacts of confirmed cases, but only a global progressive transmission-reducing intervention, the increase in contact-tracing as the number of confirmed cases increases, can be mathematically close to the dynamic increase in efficacy of control (*β*(*t*)), studied here in terms of one of the modes of control. In contrast, the social distancing alone, could be approximated to have contributed mainly a constant fixed effect on transmission reduction.

Model predictions still remain uncertain, thus it is very critical to maintain in place and strengthen the control measures to minimize transmission. As new data are arriving over the next days and weeks, with more tests to be performed, these initial predictions can be verified and updated.

The framework, however, as it stands provides a general tool to explore and understand dynamics of Covid-19 in other settings. While case-fatality-ratio here and probability of symptoms is assumed the same for the entire population, variation between age-classes or other strata of the population exist (4; 5) and could be implemented in model extensions or applications to other settings.

## Data Availability

All data are publicly available, provided by the WHO.

https://ourworldindata.org/coronavirus

## Supplementary figures

### Result of fitting the data 9-28 March with the constant control model

**Figure S1:**
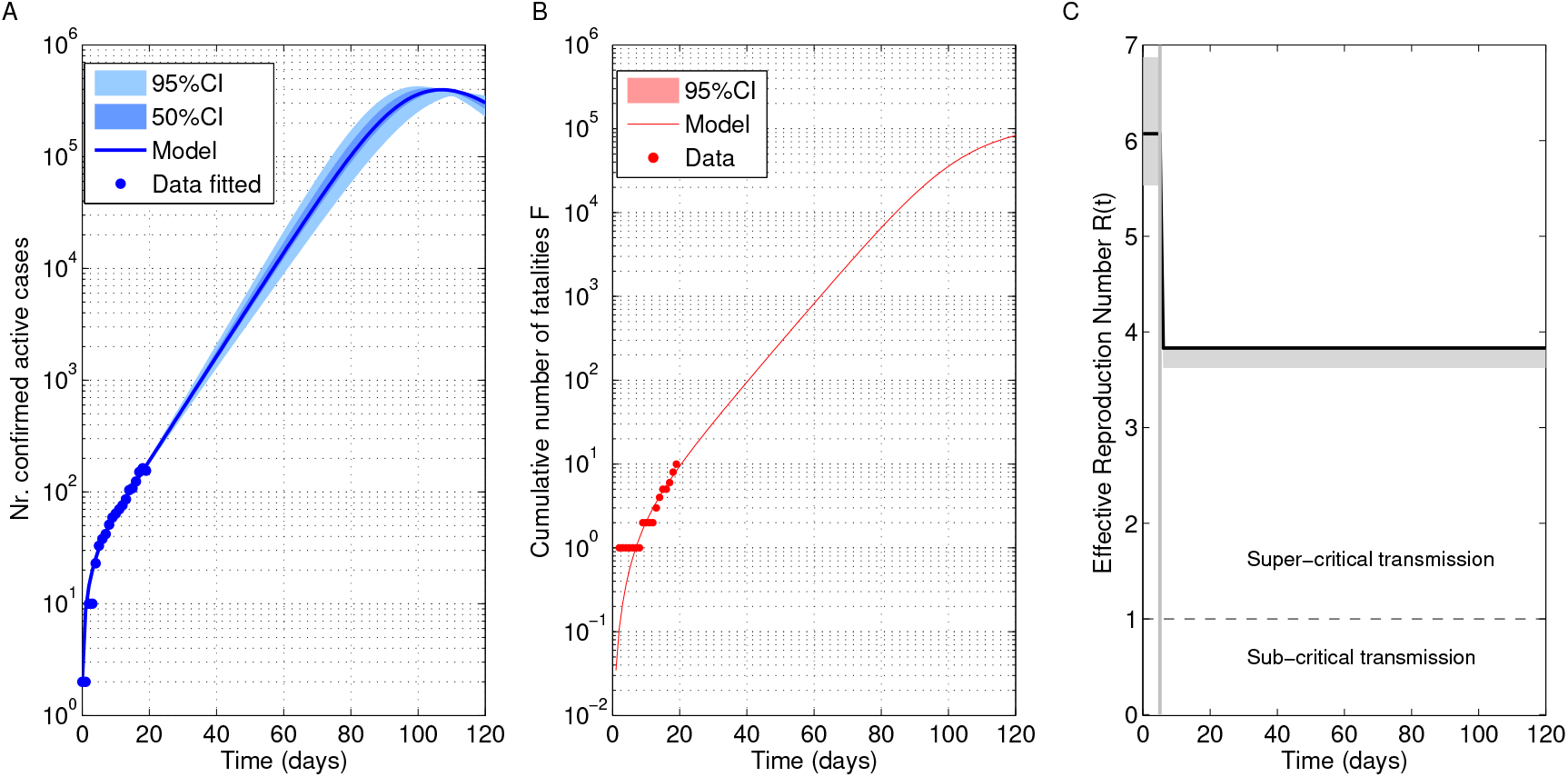
Model fit assuming the constant control model (*T*_*in*_ = 5) for data March 9-28.

**Figure S2:**
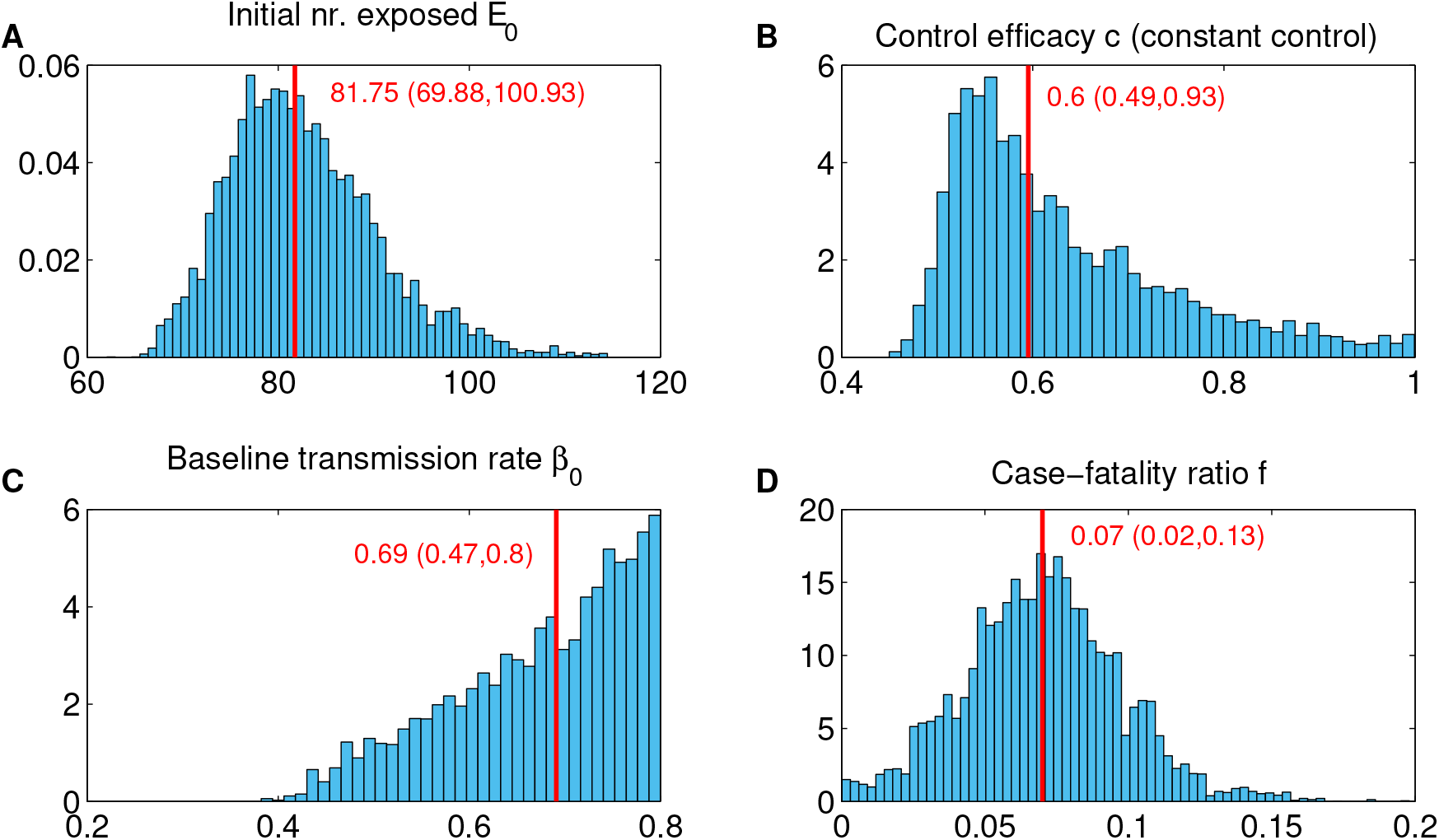
Parameter posterior distributions for model assuming constant control (*T*_*in*_ = 5) based of fitting data March 9-28.

**Figure S3:**
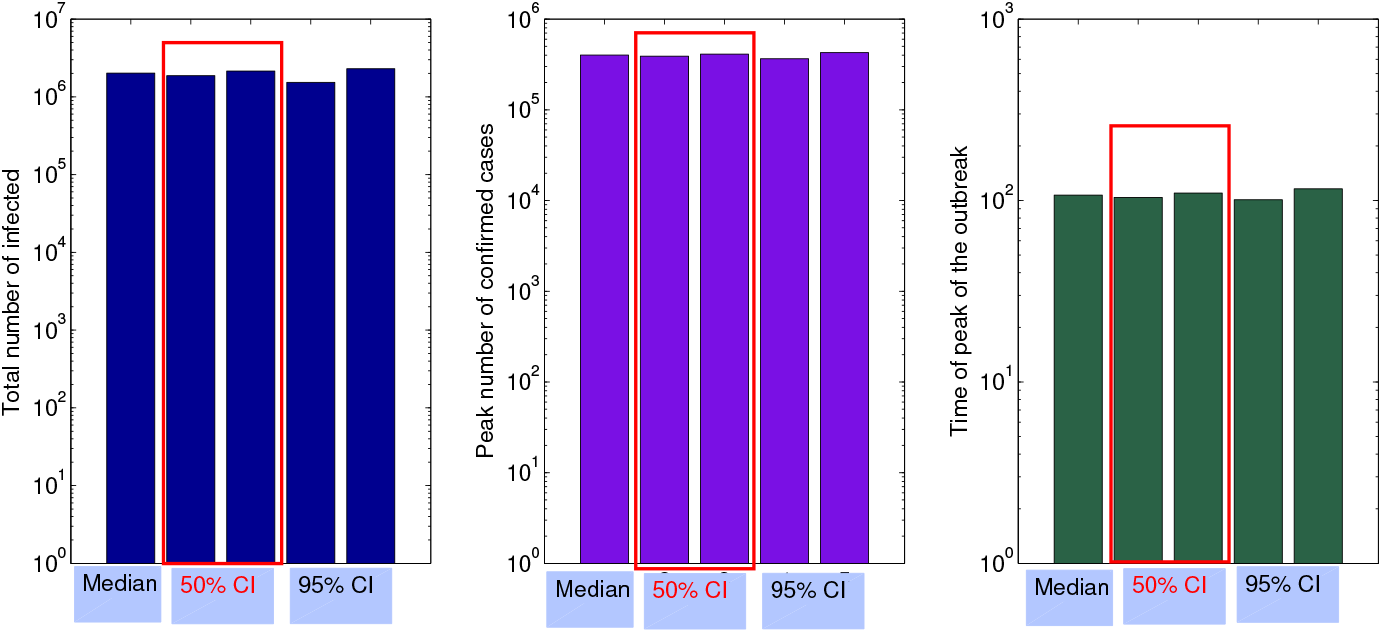
Quantitative projections with best-fitting constant control model estimates, based on model fit to March 9-28 data. A. Total number of infected. B. Peak number of confirmed (symptomatic) cases. C. Timing of the peak (number of days after March 9).

### Result of fitting the dynamic control model to data 9-31 March

**Figure S4:**
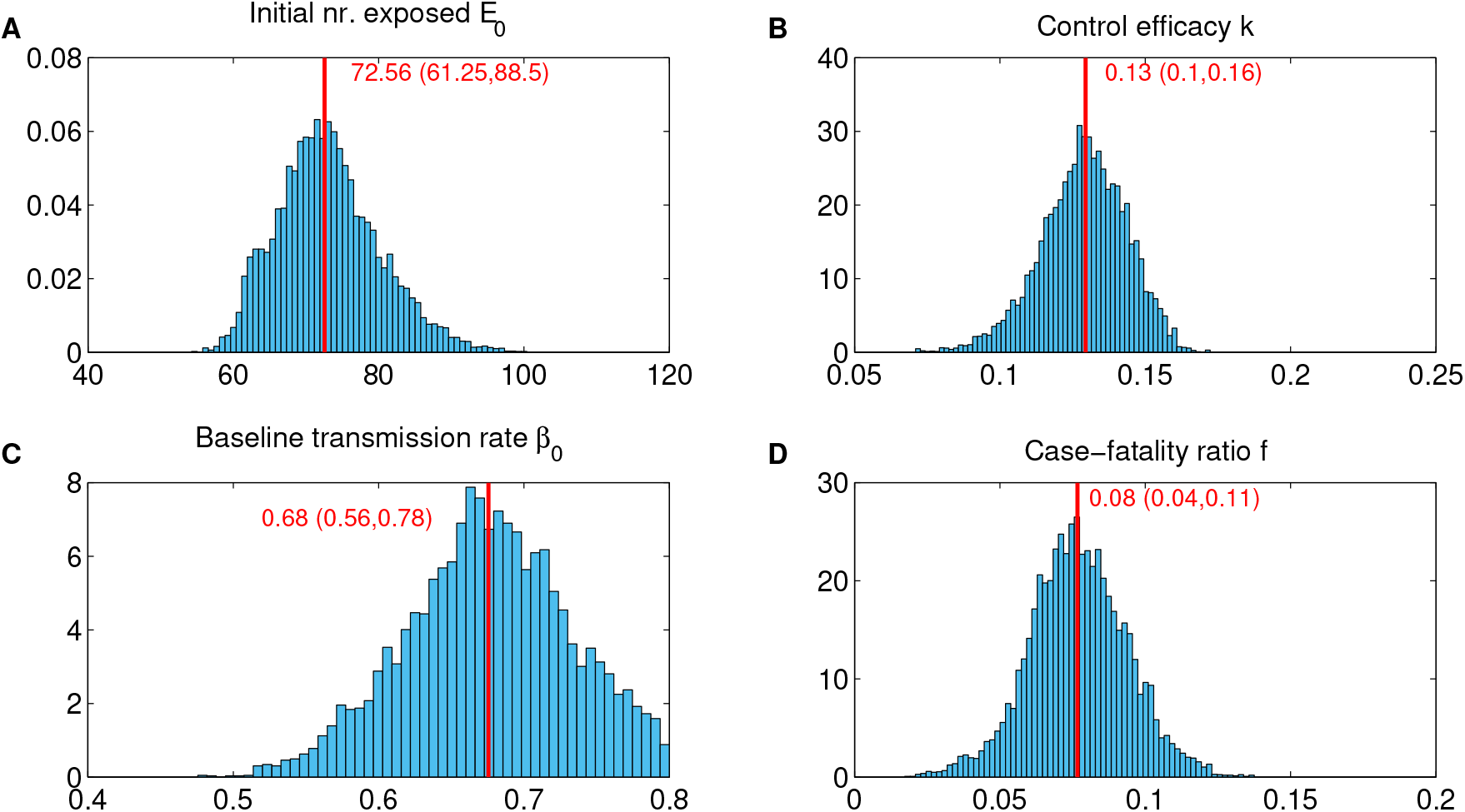
Parameter posterior distributions for dynamic control model (starting at *T*_*in*_ = 5), based of fitting data March 9-31.

**Figure S5:**
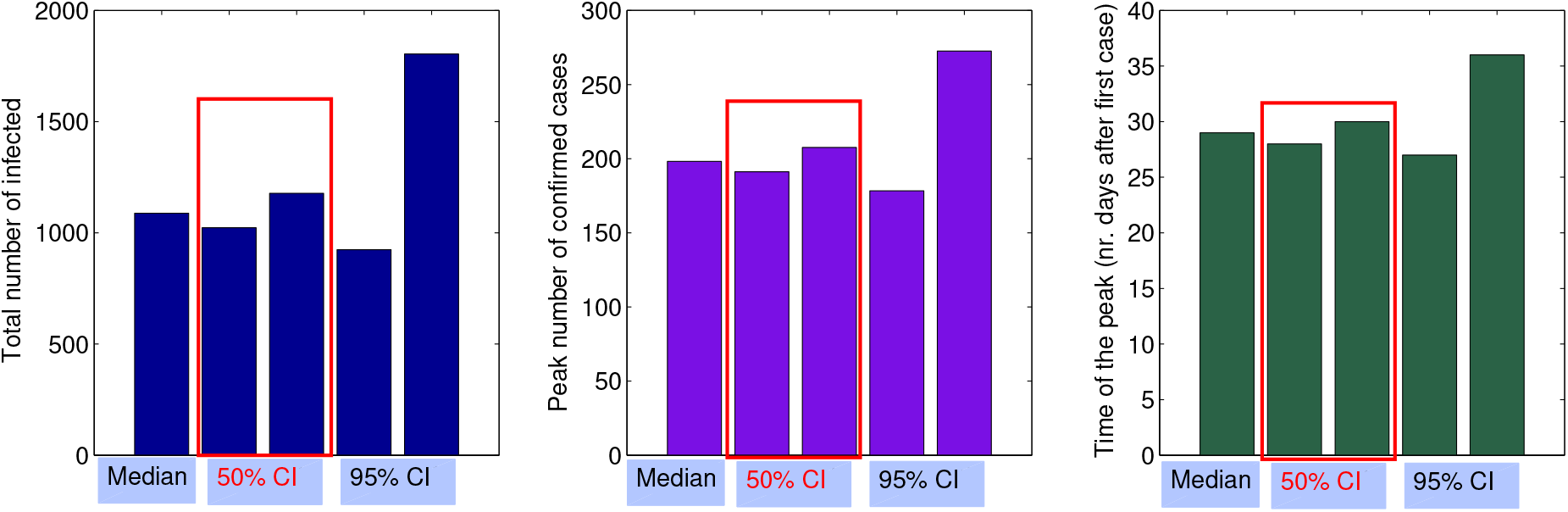
Quantitative projections with best-fitting dynamic control model estimates (onset of control assumed at *T*_*in*_ = 5), based on model fit to March 9-31 data.A. Total number of infected. B. Peak number of confirmed (symptomatic) cases. C. Timing of the peak (number of days after March 9).

## Notes

### Competing Interest Statement

The authors have declared no competing interest.

### Funding Statement

No specific funding was received for this work.

